# Biological and Disease Hallmarks of Alzheimer’s Disease Defined by Alzheimer’s Disease Genes

**DOI:** 10.1101/2022.07.16.22277709

**Authors:** Shin Murakami, Patricia Lacayo

## Abstract

An increasing number of genes associated with Alzheimer’s disease (AD) have been reported. However, despite previous studies, there is a lack of overview of the genetic relationship between AD and age-related comorbidities, such as hypertension, myocardial infarction, and diabetes, among others. Previously, we used Reactome analysis in conjunction with the genes that are associated with AD (AD genes) to identify both the biological pathways and the neurological diseases involved. Here we provide systematic updates on the genetic and disease hallmarks defined by AD genes. There are a total of 11 pathways (defined as genetic biological hallmarks) including 6 existing hallmarks and 5 newly updated hallmarks, the latter of which are developmental biology (axon and adipose development), gene expression (RNA transcription), metabolism of proteins (Amyloid formation, regulation of IGF-1/Insulin, and small ubiquitin-like modifiers/SUMOs), metabolism of RNA (mitochondrial tRNA and rRNA processing), and signal transduction (ErbB, NOTCH and p75 NTR death signaling). The AD genes further identified 20 diverse diseases that we define as disease hallmarks including: (1) existing hallmarks, including neurological diseases (Alzheimer’s disease, Amyotrophic Lateral Sclerosis, Parkinson’s Disease, and Schizophrenia); (2) as well as newly identified hallmarks, including Type 2 Diabetes, Cardiovascular diseases (Myocardial Infarction, Heart Disease, Hypertension, Cardiovascular system disease, Vascular Disease), Quantitative trait loci (lipoprotein and body mass index), Cancer (Breast cancer, Colorectal cancer, Prostate cancer, and Lung cancer); (3) and other health conditions; note that cancers reportedly have an inverse relation with AD. We previously suggested that a single gene is associated with multiple neurological diseases, and we are further extending the finding that AD genes are associated with common age-related comorbidities and others. This study indicates that heterogeneity of Alzheimer’s disease predicts complex clinical presentations in people living with AD. Taken together, the genes define AD as a part of age-related comorbidities with shared biological mechanisms and may raise awareness of a healthy lifestyle as potential prevention and treatment of the comorbidities.

## Introduction

Alzheimer’s disease is the major cause of dementia. According to the Centers for Disease Control and Prevention (CDC), 5.8 million Americans were living with AD in 2020 (CDC, 2018). Pathological characteristics of AD include diffuse and neuritic plaques characterized by amyloid plaques and neurofibrillary tangles (Vaz and Silvestre, 2020; Sherva R, Kowall, 2022). Despite these pathological characteristics, the brain pathology and progression of AD are clinically heterogeneous and thus, a clinically complex disease (Ferrari and Sorbi, 2021). Therefore, AD can be classified as late-onset (LOAD), early-onset (EOAD), and autosomal dominant forms of which LOAD is the most frequent.

AD is also highly heritable and genetically heterogeneous (Vahdati Nia *et al*., 2017; Sherva R, Kowall, 2022). Linkage analysis, genome-wide association studies and candidate gene studies have identified Alzheimer’s disease genes (AD genes). Of the 680 AD genes that are reported in the Alzgene database (Alzgene.org), 356 genes were found to be associated with AD (Vahdati Nia *et al*., 2017). Four genes are known to cause AD (APP, PSEN1 and PSEN2) or to be a risk factor (ApoE4). Based on the AD genes, a previous study identified biological Reactome pathways as biological hallmarks (Vahdati Nia *et al*., 2017). Another important finding was that AD genes are associated with 5 neurological diseases, suggesting a single gene alteration can be associated with multiple forms of neurological diseases. Here we updated and organized the biological hallmarks as well as disease hallmarks. Surprisingly, the results suggest more diverse biological hallmarks and include not only neurological diseases but also common age-related diseases, which we summarize in this study.

## Method

The method and the AD genes have been described earlier (Vahdati Nia *et al*., 2017). We used 356 AD genes. We used the updated Reactome pathway knowledgebase 2022 (reactome.org) (Gillespie *et al*., 2022) and another knowledgebase, GeneAnalytics (geneanalytics.genecards.org) (Ben-Ari *et al*., 2016). The Reactome pathways were set to a threshold of p-value ≤ 1.00E-05. To eliminate redundancies, we categorized the pathways into a spectrum ranging from general to specific: general Reactome pathways (general hallmarks), more specific pathways (more specific hallmarks), and specific pathways (specific hallmarks). For example, a Reactome analysis output in the order from general to specific was: Transport of small molecules (as general hallmarks) → Plasma lipoprotein assembly, remodelling, and clearance (as more specific hallmarks) → Plasma lipoprotein clearance (as specific hallmarks). The Reactome results of the top detected hits of AD genes include “Plasma lipoprotein clearance” and “Plasma lipoprotein assembly, remodelling, and clearance” and thus they were combined to more specific hallmarks as “Plasma lipoprotein assembly, remodelling, and clearance.” Similarly, all the redundant hits are combined and summarized.

We identified disease hallmarks, using the knowledgebase, GeneAnalytics (geneanalytics.genecards.org) (Ben-Ari *et al*., 2016) (Accessed on June 28, 2022). The knowledgebase uses a total of 74 databases. Of them, we used the results from 72 databases (Supplementary table1), excluding the results from 2 databases with potential reliability issues (Wikipathways and Wikipedia). The knowledgebase shows a range of p-values. We used the disease hits from the high tier with p-value ≤ 0.0001. The search provided the outcome as matched genes and the total genes, the latter of which included those genetically associated plus those differentially expressed in the database and thus it covers more genes than AD genes. Each disease was ranked based on the score obtained, which is based on (1) matched detected gene hits per total genes specific to each condition/quantitative trait locus; (2) the quality and the type of differentially expressed genes, genetic association and others; more details are described in the GeneAnalytics site above.

## Results

We updated specific biological pathways, using the latest Reactome knowledgebase analysis (Method). A total of 50 updated pathways were identified with a threshold of p-value less than 1.00E-05 (Supplementary Table 2). Table 1 displays the top 10 hits sorted based on their p-value. We further eliminated the redundancies among the total 50 pathways. This process generated 11 general pathways defined as general biological hallmarks and 20 more specific pathways as defined as more specific biological hallmarks (Method). Of the 11 pathways, 5 general biological pathways are existing hallmarks reported in the earlier study (Vahdati Nia *et al*., 2017).

**Table 1.**
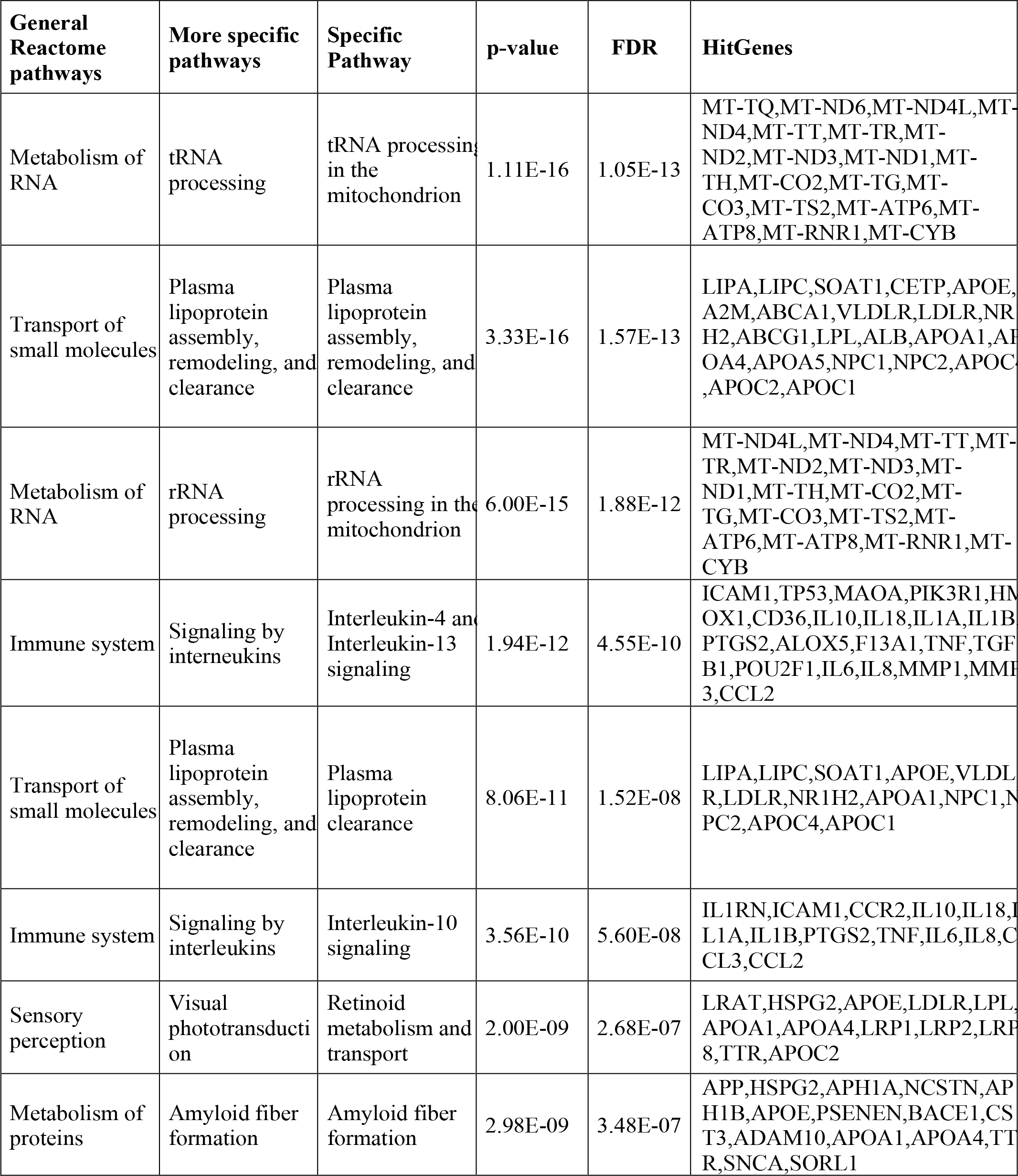
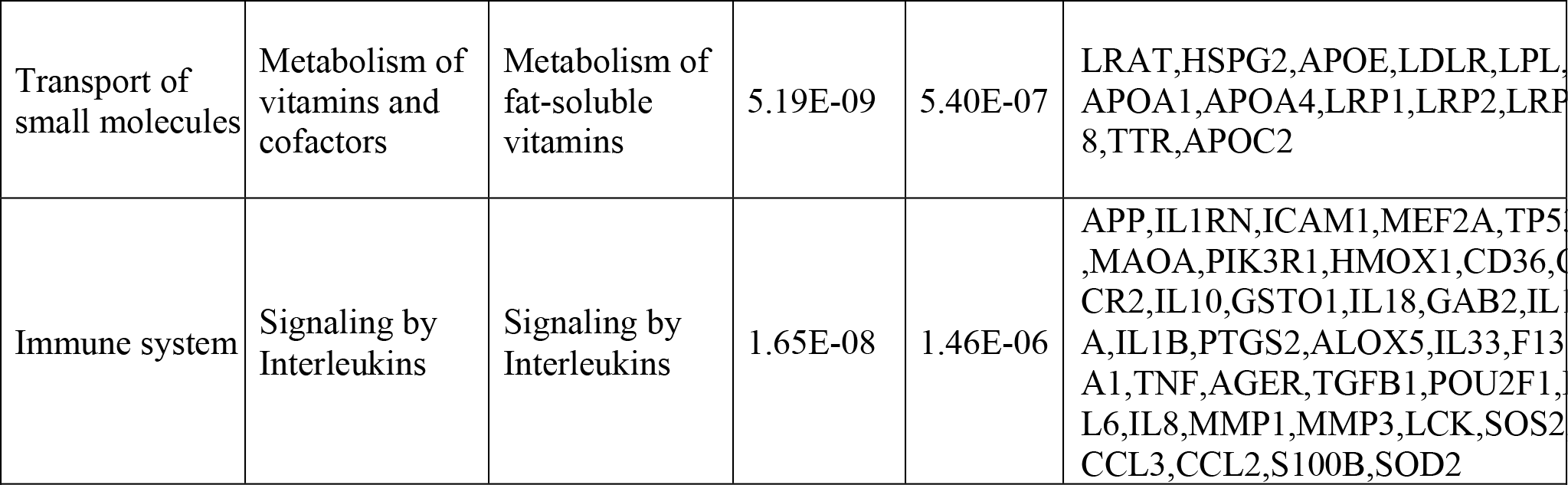
Updated top 10 Reactome pathways. The Reactome analysis updated the biological pathways that we define as biological hallmarks (Accessed on June 28, 2022). FDR (False detection rate).

Figure 1 summarizes 11 general pathways defined as general biological hallmarks and 20 more specific pathways (defined as more specific biological hallmarks). The 11 general hallmarks (with keywords) are in alphabetical order (asterisks[*] indicate newly identified hallmarks):

1. Developmental Biology (axon and adipose development)*
2. Extracellular matrix organization (protein degradation)
3. Gene expression (RNA polymerase II transcription)*
4. Hemostasis (platelet regulations)
5. Immune System (interleukins)
6. Metabolism (lipoproteins, fat-soluble vitamins, eicosanoids/steroids)
7. Metabolism of proteins (Amyloid formation, regulation of IGF-1/Insulin, and small ubiquitin-like modifiers/SUMOs)*
8. Metabolism of RNA (mitochondrial tRNA and rRNA processing)*
9. Sensory perception (retinoids)
10. Signal Transduction (ErbB, NOTCH and p75 NTR death signaling)*
11. Transport of small molecules (lipoproteins)

Of the 11 pathway hallmarks, 5 were newly updated hallmarks. “Developmental Biology” include a subcategory in axon and adipose development. Axon development was divided into EPH-Ephrin signaling and EPH-ephrin mediated repulsion of cells; the adipose development is from transcriptional regulation of white adipocyte differentiation. “Gene expression” includes RNA Polymerase II Transcription. Although “Metabolism” is an existing hallmark, eicosanoid/steroid is a new subcategory, featuring the synthesis of 5-eicosatetraenoic acids. Notably, “Metabolism of proteins” consists of the pathway directly involved in AD (amyloid formation) and two new pathways, regulation of endocrines and lifespans (regulation of IGF-1/insulin) and protein turnover (small ubiquitin-like modifiers/SUMOs). Another hallmark includes a new category “Metabolism of RNA”, which is mitochondrial tRNA and rRNA processing in mitochondria. Lastly, “Signal transduction” includes nuclear receptor signaling, including NR1H2 and NR1H3-mediated signaling, ErbB signaling and p75 NTR death signaling, while NOTCH signaling was reported previously (Vahdati Nia *et al*., 2017).

**Figure 1.**
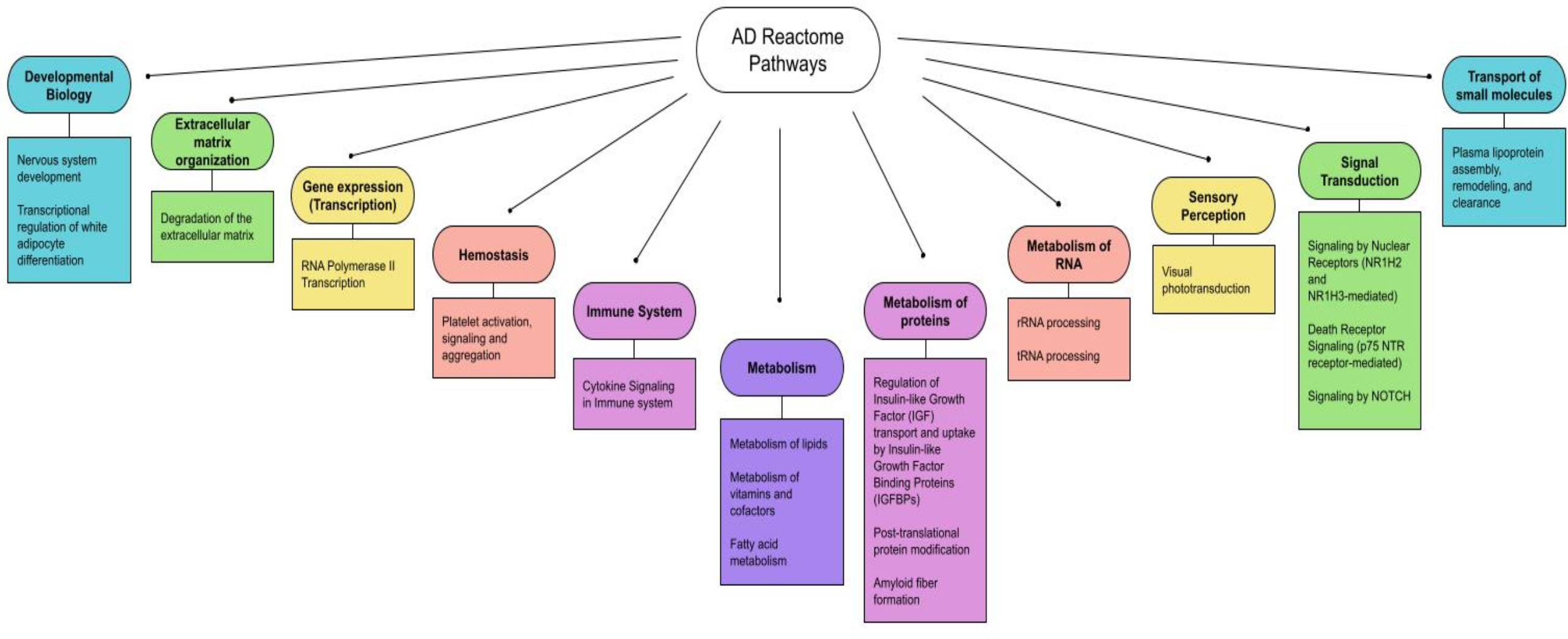
Updated biological hallmarks. We extended the list indicated by Table 1, using the threshold (p-value < 1.00E-05) and summarized the 11 general biological hallmarks (circle) and more specific hallmarks (square).

We further identified diseases associated with AD genes, using the GeneAnalytics knowledgebase (Method). The knowledgebase ranks the association based on 74 databases by tiers (Method). The detected gene hits of the top 20 diseases are summarized in Table 2 (p-value ≤ 0.0001). Disease hallmarks are summarized in Figure 2. Based on the types of diseases, the AD genes can be classified as: (1) genes specific to neurological diseases; (2) genes more general to common age-related diseases; and (3) genes general to others. The first group of diseases was neurological diseases (Alzheimer’s disease, General and peripheral nervous system disease, Amyotrophic Lateral Sclerosis, Parkinson’s Disease, and Schizophrenia), which were reported previously (Vahdati Nia *et al*., 2017). 112 AD genes were matched out of 836 AD1 genes that are either genetically associated or differentially expressed. AD1 is a specific type of AD caused by mutations in the APP gene, a source of beta-amyloid. Of a total of 356 AD genes, the gene hits of 112 accounts for 31.4% of AD genes (112 out of 356 AD genes). The second group of diseases are common age-related diseases. They include type 2 diabetes, cardiovascular diseases (myocardial infarction, heart disease, hypertension, cardiovascular system disease, and vascular disease), cancer (breast cancer, colorectal cancer, prostate cancer, and lung cancer) and others (osteoporosis). Thus, alterations in AD genes are associated with age-related comorbidities in addition to AD. The third group introduces other conditions including cystic fibrosis and quantitative trait loci (lipoprotein and body mass index). Surprisingly, cystic fibrosis is included in the disease hit by AD genes.

**Table 2.** Diverse disease hallmarks are associated with AD genes. The top 20 diseases in the high tier (p-value equal or less than 0.0001) are listed. The AD gene sets in Table 1 are used to identify diseases using all 356 AD genes (Vahdati Nia *et al*., 2017) and the web-based search using GeneAnalytics (Accessed on June 28, 2022). *, The number of AD gene hits (total genes classified in each group of the health conditions/Loci).

| General criteria | Health conditions/loci | # Matched gene hits (Total genes)* |
| --- | --- | --- |
| Neurological | Alzheimer Disease, Familial, 1 | 112 (836) |
| Neurological | Nervous System Disease | 91 (897) |
| Diabetes | Type 2 Diabetes Mellitus | 84 (555) |
| Cardiovascular | Myocardial Infarction | 68 (302) |
| QTL* | Body Mass Index Quantitative Trait Locus 11 | 76 (819) |
| Cancer | Breast Cancer | 73 (1447) |
| Cardiovascular | Heart Disease | 61 (366) |
| Cardiovascular | Hypertension, Essential | 59 (457) |
| Cancer | Colorectal Cancer | 70 (1492) |
| Neurological | Amyotrophic Lateral Sclerosis 1 | 55 (579) |
| Cardiovascular | Cardiovascular System Disease | 51 (192) |
| Neurological | Parkinson Disease, Late-Onset | 52 (387) |
| Other | Lipoprotein Quantitative Trait Locus | 49 (248) |
| Cardiovascular | Vascular Disease | 44 (149) |
| Neurological | Schizophrenia | 48 (464) |
| Neurological | Peripheral Nervous System Disease | 46 (369) |
| Cancer | Prostate Cancer | 56 (1121) |
| Other | Osteoporosis | 44 (308) |
| Cancer | Lung Cancer | 50 (1218) |
| Other | Cystic Fibrosis | 40 (333) |

**Figure 2.**
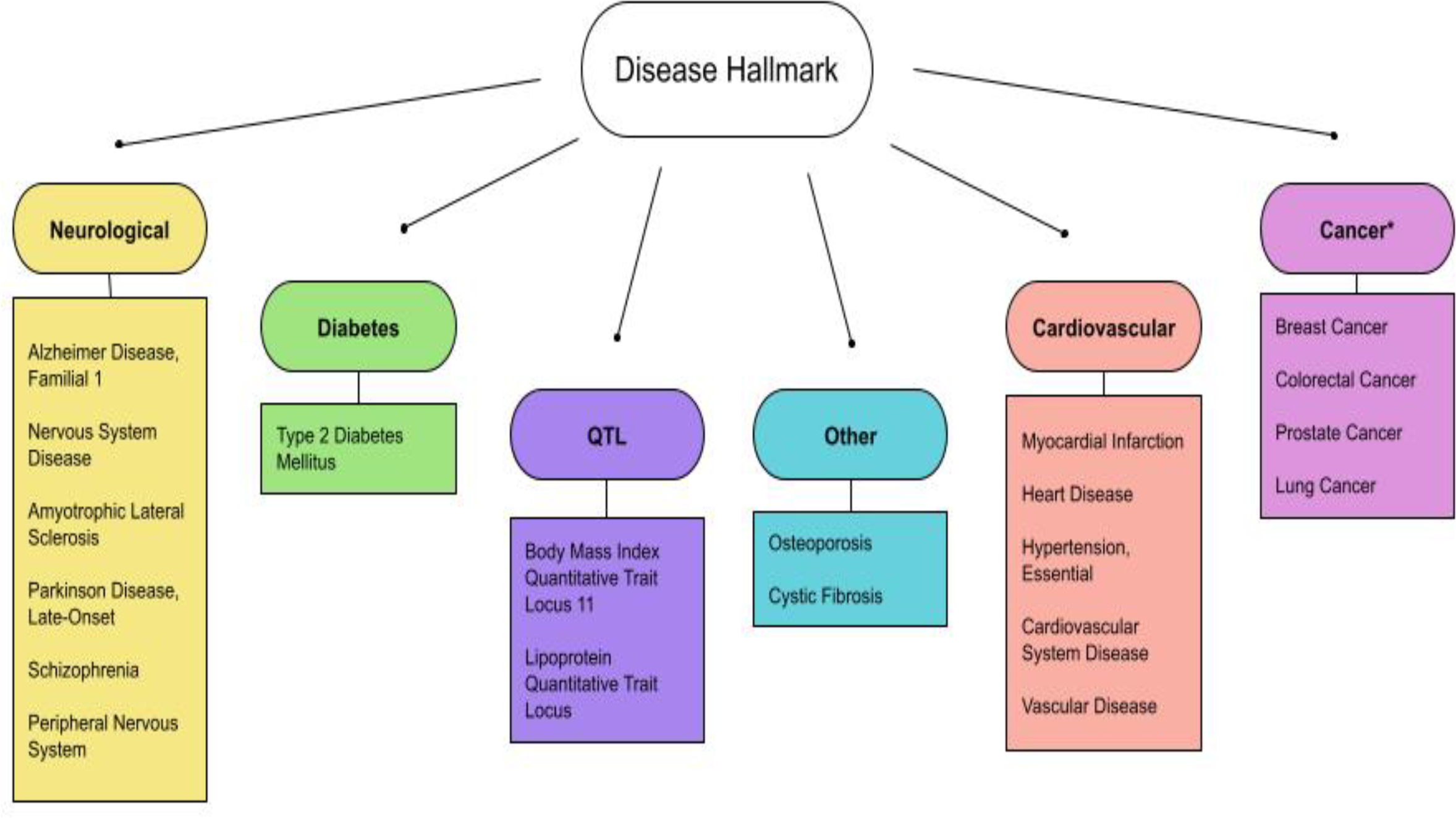
Updated disease hallmarks. The diseases listed in Table 2 are organized and grouped into 6 general disease criteria. They are further classified into: (1) neurological diseases (Alzheimer’s Disease, Familial, 1, Amyotrophic Lateral Sclerosis 1, Parkinson Disease, Late-Onset, Schizophrenia, Peripheral Nervous System Disease, Nervous System Disease, others); (2) common age-related diseases (Diabetes, Cardiovascular Disease, Cancer and Osteoporosis); and (3) other diseases (Quantitative Trait Loci and Cystic fibrosis). Note that cancers are reported to show an inverse relationship with AD genes (indicated by asterisks).

## Discussion

This study updated genetic hallmarks for both biological Reactome pathways and those for diseases. We identified 11 general biological pathways, which included 5 existing pathways and 6 new pathways. The existing biological pathways include: The “immune system” unfolds pathways involving interleukin-4, 10, and 13 which are involved in the pathology of a wide variety of age-related diseases such as cardiovascular diseases, diabetes and cancers. Similarly, “Metabolism” includes lipoprotein dysregulations relevant to dyslipidemia, and cardiovascular diseases, among others. The previous version of Reactome knowledgebase classified retinoid metabolism as “Metabolism”, yet the renewed 2022 version classified it as “Sensory perception”. Retinoids or vitamin A are part of fat-soluble vitamins. Thus, we included “Sensory perception (retinoids)” back to the existing pathway of “Metabolism (fat-soluble vitamins)”.

Of the 6 new biological pathways, “Metabolism of RNA” includes mitochondrial tRNA and rRNA processing in mitochondria. It may be consistent with mitochondrial deletions known to occur during aging, which may cause mitochondrial deficits (Swerdlow *et al*., 2018; Jang *et al*., 2018). Another category “gene expression” includes RNA Polymerase II Transcription, which is consistent with common age-related transcriptional changes (Stegeman and Weake, 2018). The category impacts the expression of a wide variety of stress response genes (Stegeman and Weake, 2018). Related to this, stress resistance is a component of life extension in model systems (Johnson *et al*., 2000; Murakami, 2007; Hamilton and Miller, 2017; Buono and Longo, 2018). Metabolism eicosanoid/steroid is a new subcategory including the synthesis of 5-eicosatetraenoic acids, which is a part of the eicosanoid pathways for lipoxygenase (LOX) and cyclo-oxygenase (COX) pathways among others. The category “Metabolism of proteins,” includes the pathway of amyloid formation, regulation of IGF-1/insulin, and small ubiquitin-like modifiers (SUMOs). Amyloid formation is directly involved in beta-amyloid plaques. The impaired insulin pathway causes diabetes, which is closely related to AD (Moreira 2012; Baglietto-Vargas *et al*., 2016). SUMOs are involved in protein turnover which is also associated with AD (Hendriks and Vertegaal, 2016). Lastly, “Signal transduction” includes nuclear receptor signaling, including NR1H2 and NR1H3-mediated signaling, ErbB cancer signaling and p75 NTR death signaling; note that NOTCH signaling was reported previously (Vahdati Nia *et al*., 2017). NR1H2 and NR1H3 are known as liver-X receptors (LXR), which regulate cholesterol metabolism. They are receptors for their ligands, oxysterols, that are generated by the oxidation of cholesterol through ROS (reactive oxygen species) and other processes (Ma and Nelson ER, 2019). ErbB and p75 NTR are involved in cell survival and death.

The diseases identified by using AD genes can be categorized into 3 major criteria (neurological diseases, common age-related diseases, and others) and broken down into 6 general disease criteria. The first disease criterion is neurological diseases which include Alzheimer’s disease, general and peripheral nervous system disease, amyotrophic lateral sclerosis, Parkinson’s disease, and schizophrenia. This group has been reported and discussed in the previous study (Vahdati Nia *et al*., 2017). The study concluded that a single gene alteration may cause multiple neurological diseases. The top hit of the disease in this study was AD1 (Alzheimer’s Disease, Familial, 1). Based on affected genes, the efforts on classifying AD have been ongoing and currently classified from AD1 to AD16. AD1 is caused by mutations in the APP genes. AD2 is associated with the ApoE4 allele. AD3 is caused by mutations in PSEN1. AD4 is caused by mutations in the PSEN2 gene. For more details, see the GTR (genetic testing registry) at the NCBI (National Center for Biotechnology Information database) (GTR, 2022, Accessed July 12, 2022). Due to the number of AD genes, the list of AD types is expected to be increased in number. The study provides a clear example of genetic and phenotypic heterogeneity. The result is consistent with the complex clinical presentations (i.e., clinical heterogeneity) of AD (Ferrari and Sorbi, 2021).

The second and third disease criteria include common age-related diseases. Age-related diseases are also seen in people as age-related comorbidities, with which two or more diseases commonly occur in a single person. The comorbidities include diabetes (type 2 diabetes), cardiovascular diseases (myocardial infarction, heart disease, hypertension, cardiovascular system disease, and vascular disease), cancer (breast cancer, colorectal cancer, prostate cancer, and lung cancer) and others (osteoporosis). It is worth noting that cancers show an inverse relation with AD (Nudelman *et al*., 2019). The observation is consistent with the previous studies that a major hypertension target, angiotensin-converting enzyme (ACE) is also involved in AD (Le *et al*., 2020 and 2021). Despite being less defined, AD may be classified as type 3 diabetes, which is a type of diabetes in the brain (Steen *et al*., 2005; Pilcher, 2006; de la Monte, 2014; Leszek *et al*., 2017). The vast majority of AD falls into LOAD, whose onset occurs starting at 65 years of age, while age-related diseases occur earlier than that. Although age-related comorbidities are known to be vulnerable to a variety of conditions, for example, COVID-19 (Antos *et al*., 2021), the straightforward interpretation of the result is that AD genes are associated with AD as well as with common age-related diseases.

It is conceptually important that the AD genes define AD as a part of age-related comorbidities with shared biological mechanisms. While the study raises the possibility that age-related diseases may lead to AD, we are more inclined to the possibility that the shared biological mechanisms may lead to AD and other age-related comorbidities. We are beginning to learn that “there is growing evidence that people who adopt healthy lifestyle habits…can lower their risk of dementia…which have been shown to prevent cancer, diabetes, and heart disease (CDC, 2020)”. Moreover, the CDC describes the broad neurological behavioral warning signs of Alzheimer’s disease, such as memory impairment, difficulty in daily tasks, and poor judgement, among others (CDC, 2020). This study further suggests that common age-related comorbidities may present early signs when AD genetics is involved. Related to this, a wide variety of clinical scenarios may be considered. For example, people living with AD gene alterations may have common age-related comorbidities and risk of AD development; people living with AD gene alterations may have AD with other common age-related comorbidities or people living with AD gene alterations may develop neurological and other conditions.

The cure for AD is still unknown. Currently, Aducanumab, a human anti-beta-amyloid antibody, is the only disease-modifying medication approved by FDA (U.S. Food and Drug Administration) (National Institute on Aging, 2021; Alzheimer’s Association, 2022). The medication requires assessment of brain beta-amyloid, which uses PET (positron emission tomography) scans or analysis of cerebrospinal fluid. As a clinical approach to AD, we present that age-related comorbidities may provide an early assessment when genetic testing is performed. We also present that the treatment options for age-related comorbidities may be effective when biological mechanisms are considered. Alternatively, the implications from the model systems may be useful for treatment options. Stress resistance confers resistance to multiple forms of stressors, such as pathogens and the toxic beta-amyloid, which is tightly associated with Alzheimer’s disease in the model systems (Florez-McClure *et al*., 2007; Machino *et al*., 2014). Multiplex stress resistance is a key to understanding the mechanism of extended lifespans and health spans (Murakami *et al*., 2003; Murakami, 2006). Additionally, stress resistance is tightly associated with life-extending interventions (Murakami, 2006) in which the molecular mechanisms are genetically characterized, for example, the insulin/IGF-1 pathways (Murphy and Hu, 2013), and serotonin pathways (Murakami and Murakami, 2007), among others; these can be assessed by semi-automated systems (Machino *et al*., 2014). The IGF-1/insulin pathways are a major regulator of lifespans (Finch and Ruvkun, 2001; Kenyon, 2010; Ewald *et al*., 2018; Zhang *et al*., 2020) and are involved in age-related memory impairment (Murakami *et al*., 2005). Similarly, the serotonin pathways regulate age-related behavioral changes, lifespans and stress resistance (Murakami and Murakami, 2007; Murakami *et al*., 2008). More details of age-related memory impairment and a related theory (middle-life crisis theory of aging) are described elsewhere (Murakami *et al*., 2011; Murakami, 2013). Taken together, we suggest that this study of revisiting AD genes provides the strength of treatment options as well as future direction. It will be a powerful way to develop a science-based tool for the long-waited diagnosis, prevention, and treatment options for AD.

## Data Availability

All data produced in the present work are contained in the manuscript

## Acknowledgement

This study was devoted to helping my friends and people living with Alzheimer’s disease. We thank Student Doctors Timothy Balmorez and Amy Sakazaki and other Murakami lab members for discussion and comments on the manuscript.

